# Associations between prenatal (poly)phenol intake and cognitive outcomes in school-age children

**DOI:** 10.64898/2026.10.08.26365020

**Authors:** Chelsey Fiecke, Jayne Bellando, Mara Whiteside, Chayla Slaton, Milan Bimali, Miaolei Bao, D. Keith Williams, Ellen van der Plas, Colin D. Kay, Aline Andres

**Author notes:** Correspondence: Chelsey Fiecke, 15 Children’s Way, Little Rock, AR 72202.

## Abstract

**Background:** (Poly)phenol intake is associated with improved cognitive function in adults, with emerging evidence suggesting benefits in children. However, the role of prenatal (poly)phenol exposures in cognitive development remains a gap in early-life nutrition research.

**Objective:** This analysis of a longitudinal cohort tested the associations between prenatal (poly)phenol intakes and cognitive outcomes in school-age children at age 5 and 8 years.

**Methods:** Participants included mother-child pairs enrolled in a prospective, longitudinal study during pregnancy and completed 5- and 8-year follow-up visits (N = 129). 3-day food records were used to estimate prenatal and child intakes of energy and 18 (poly)phenol subclasses using a database developed from Phenol-Explorer (v3.6). The analytical sample included children (N = 111) with completed assessments for academic achievement, language skills, and intelligence quotient (IQ) scores (Mean = 100, SD = 15). Regression models estimated associations between (poly)phenol intakes and cognitive outcomes, adjusted for breastfeeding duration, maternal IQ, pre-pregnancy BMI, gestational age, and infant sex.

**Results:** Average prenatal total (poly)phenol intake was 716 ± 383 mg/d. Prenatal (poly)phenol intakes were not associated with child academic achievement scores. Higher prenatal flavone intakes were associated with higher language (β: 4.36, p = 0.045) and IQ (β: 9.43, p = 0.002) scores. Higher flavonol (β: −4.67, p = 0.022) or hydroxybenzoic acid intakes (β: −3.88, p = 0.026) were associated with lower language scores. Major sources of flavones and lignans included coffee, orange juice, and herbal tea. Coffee, cranberry, black tea, okra, and tomato sauce provided flavonols and hydroxybenzoic acids.

**Conclusions:** Associations between prenatal (poly)phenol intake and child cognitive outcomes were mixed and differed by subclass. Further investigation is needed to explore the role of co-occurring nutrients and other dietary components. These findings suggest that (poly)phenol subclasses derived from healthier plant-based foods, including citrus fruit and herbal tea, may associate with improved child cognitive outcomes.

**Clinical Trial Registry number:** NCT01131117, https://clinicaltrials.gov/study/NCT01131117; NCT03108001, https://clinicaltrials.gov/study/NCT03108001

## Introduction

The first 1,000 days of life (conception to 2 years of age) represents a critical period of neurodevelopment that impacts infancy, childhood, and adulthood^1–3^. It is well-established that early-life nutrition exposures during this period, spanning pregnancy^4, 5^, infant and toddler feeding practices^6–8^, and childhood diets^9^, have long-term implications for offspring cognitive function. Fetal neurodevelopment is particularly nutrient-sensitive due to neurogenesis, neuronal migration, synaptogenesis, and myelination occurring during this time period^10^. Well-established examples of nutrients with known roles in offspring brain development include long chain omega-3 polyunsaturated fatty acids (docosahexaenoic acid [DHA])^4^, folate^11^, iron^5^, iodine^12^, zinc^13^, vitamin B_12_ ^14^, vitamin K^15^, and choline^4^. Beyond micronutrients, biologically active plant-based compounds, such as (poly)phenols and carotenoids, are associated with improved diet quality^16, 17^ and are increasingly recognized for potential impacts on cognitive function in children^18–20^ and adults^21, 22^. However, prior studies isolating the impacts of (poly)phenols on child cognitive health are limited to two acute blueberry dosing studies in school-age children (8-10 years^20^ and 7-10 years of age^19^) with small sample sizes (N = 14 and N = 21, respectively).

Unlike intakes of other nutrients, which are single compounds that have a long history of evaluation during pregnancy, (poly)phenols represent a broad class of over 8,000 individual compounds^23^ categorized into 5 major classes (flavonoids, phenolic acids, lignans, stilbenes, ellagitannins) and 18 subclasses, with limited data on intakes during pregnancy. Efforts to characterize prenatal (poly)phenol intakes include international cohorts based in Brazil^24^, China^25, 26^, and Spain^27^ (average intake: 1,278 mg/d). To our knowledge, prenatal (poly)phenol intakes have yet to be characterized in a U.S.-based cohort, due in part to the systematic exclusion of pregnant women from population-level estimates of (poly)phenol intakes^28, 29^. Data on (poly)phenol intakes in children in the U.S. is also limited, despite efforts to characterize (poly)phenol intakes internationally (average intake for children ≥ 4 years of age: 612 mg/d)^30–33^. However, prior efforts to estimate prenatal and child intakes have focused primarily on total (poly)phenol intakes without subclass-level characterization and considerable heterogeneity exists in methodologies (e.g., (poly)phenol content database selection, estimation of (poly)phenol intakes based on the non-specific Folin-Ciocalteu assay vs. chromatographic methods)^24, 30, 32, 34^. In addition, (poly)phenol intakes in pregnant mothers and their children have yet to be evaluated using prospective, longitudinal designs.

Emerging evidence supports offspring cognitive health benefits of consuming (poly)phenol-rich dietary patterns (e.g., fruit and vegetable consumption, Mediterranean diet adherence) during pregnancy. Higher prenatal fruit intakes are associated with increased cognitive and adaptive behavior scores at age 1 year^35^ and reduced risk of developmental delay at age 2 years^36^. Prenatal vegetable intakes are also associated with reduced risk of developmental delay at age 2 years^36^. Similarly, Mediterranean diet adherence during pregnancy is associated with reduced risk of communication development delay at age 1 year^37^. With the exception of Yonezawa et al.^36^, evaluations of associations between prenatal intakes of (poly)phenol-rich dietary patterns and child cognitive outcomes rarely account for child dietary intake beyond breastfeeding status or duration. Despite emerging evidence that consuming (poly)phenol-rich dietary patterns during pregnancy associates with improved child cognitive outcomes, prenatal (poly)phenol intake has yet to be isolated as an independent predictor of improved offspring cognitive health.

This is the first study to estimate (poly)phenol intake in both pregnant mothers and their children, filling a critical gap in early-life nutrition research. In this analysis, we aimed to characterize (poly)phenol intake during pregnancy and childhood for mother-child dyads enrolled in a longitudinal cohort and investigate associations of prenatal (poly)phenol intake with child cognitive outcomes at age 5 and 8 years. The findings lay the foundations to design targeted interventions to test whether (poly)phenol-rich diets during key developmental windows can improve children’s cognitive health.

## Methods

### Participants

Participants were mothers and their children who were enrolled in a prospective, longitudinal cohort study at the Arkansas Children’s Nutrition Center in Little Rock, Arkansas (NCT01131117)^38^ and a follow-up study that recalled the children of the parent cohort at age 5 and 8 years (NCT03108001)^39^. Women were enrolled prior to 10 weeks of gestation. Details related to the study design, inclusion and exclusion criteria, have been described previously^40, 41^. Briefly, women were excluded from the study if they had pre-existing conditions (e.g., diabetes mellitus, hypertension); pregnancy complications; used medications during pregnancy that influence fetal growth; or used recreational drugs, tobacco, or alcohol.

Participants who completed the 24-month visit (N = 194) of the parent cohort were contacted for participation in the follow-up study at age 5 and 8 years. A total of 129 children were enrolled in the follow-up study, with 103 children completing the 5-year visit (80%) and 91 completing the 8-year visit (71%). Participants in the follow-up study that had at least one cognitive assessment completed at age 5 or 8 years and at least one 3d food record per trimester during pregnancy were included for analysis (5y: N = 90; 8y: N = 80). The entire analytical sample consisted of 111 mother-child dyads. A participant flow chart for is shown in **Figure 1**. The study procedures were approved by the Institutional Review Board of the University of Arkansas for Medical Sciences.

**Figure 1:**
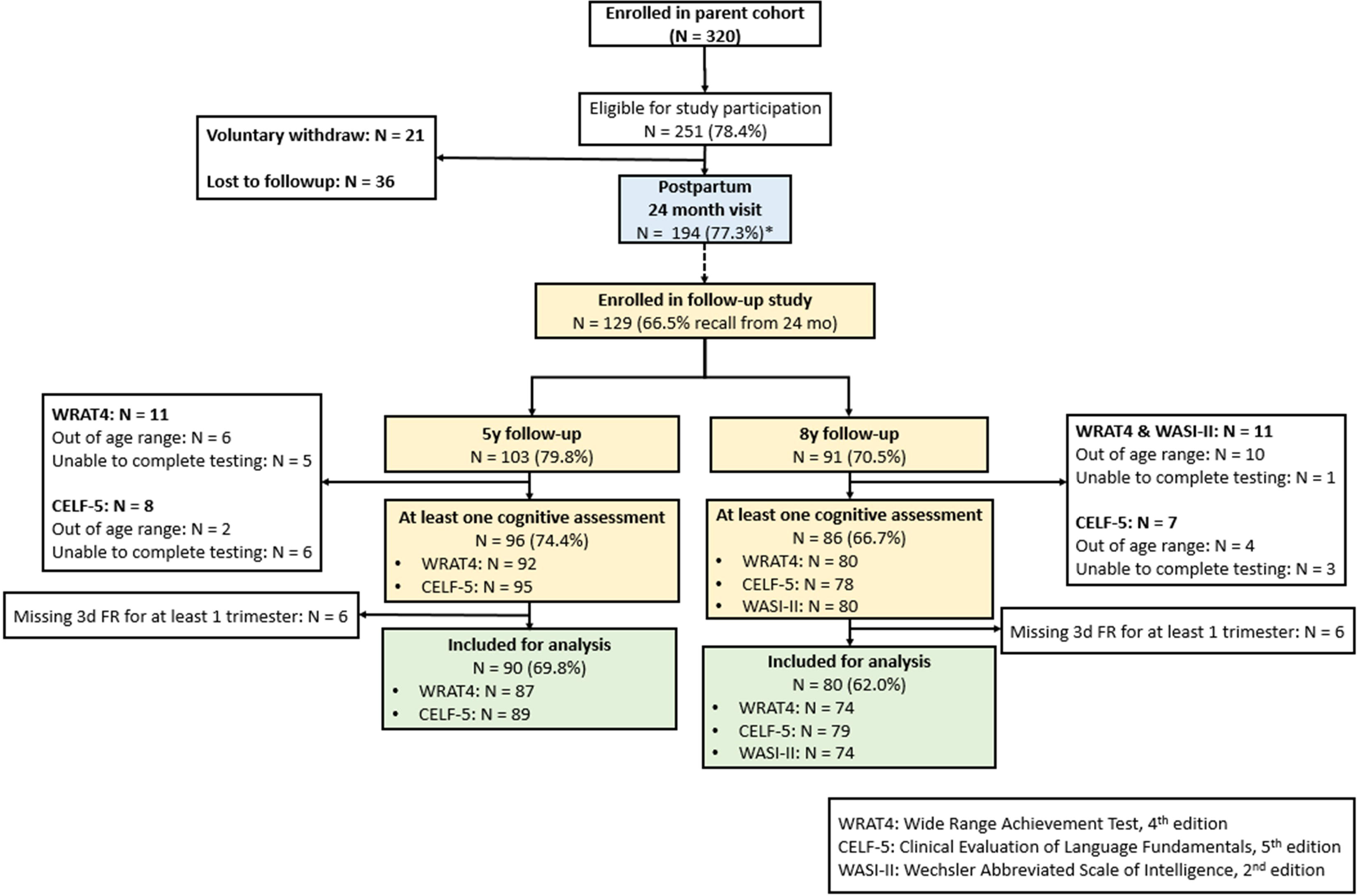
STROBE flow diagram of participants through the parent cohort (pregnancy through 24 months postpartum and the follow-up study (visits at 5 and 8 years) included for analysis (N = 111). *Percentage represents the participants out of all eligible participants. FR: food record.

### Dietary assessment

Maternal and child dietary intake were evaluated using 3-day food records analyzed using the Nutrition Data System for Research software (NDSR, Nutrition Coordinating Center [NCC], University of Minnesota, MN). Participants were instructed to record food, beverage, supplement, and medication intake for 2 weekdays and 1 weekend day. Food records were completed at <10, 12, 18, 24, 30, and 36 weeks of gestation for the mother and at age 5 and 8 years for the child. Average intakes of energy (kcal/d) and individual food items (g/d) were calculated for each visit.

### Estimation of (poly)phenol intake

Maternal and child (poly)phenol intakes were estimated using a database developed from Phenol-Explorer (version 3.6)^42^. **Figure 2** shows linkages between components of the databases.

**Figure 2:**
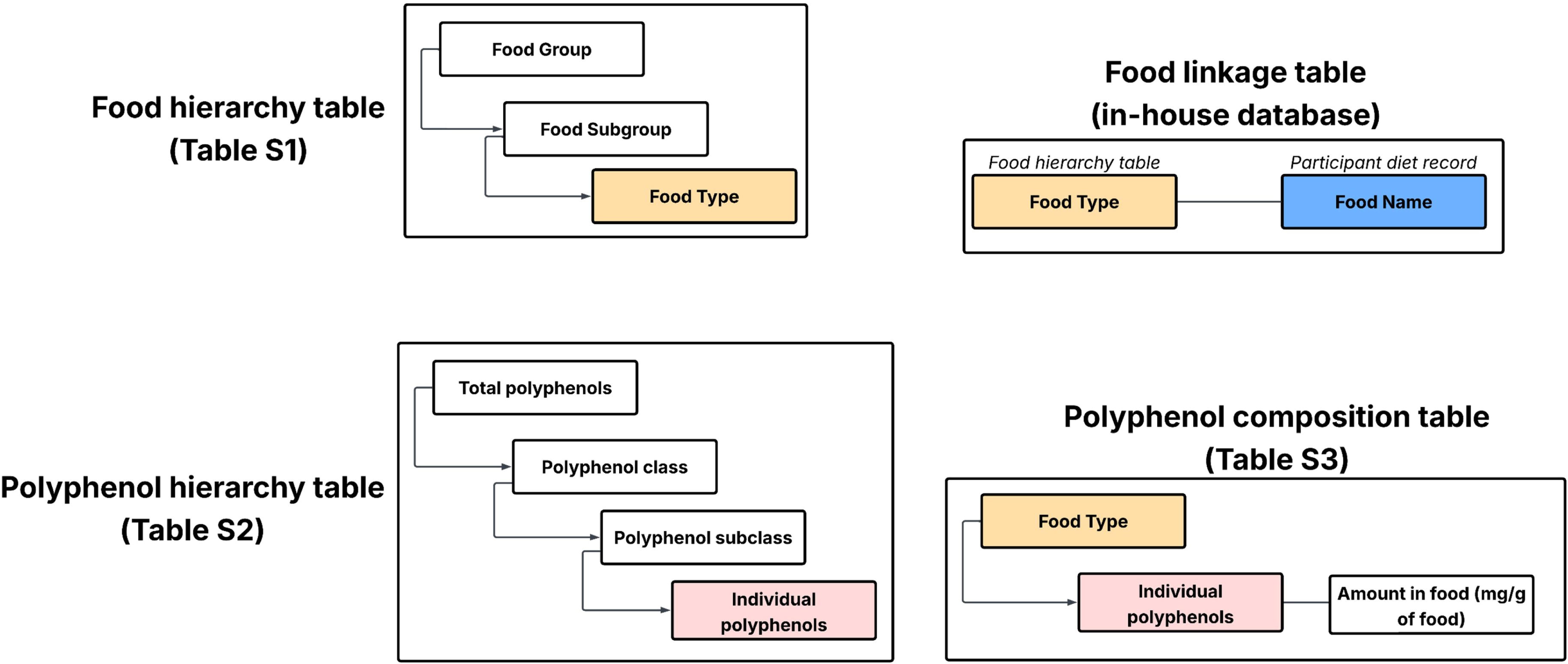
Linkages between components of the database developed to estimate (poly)phenol intakes from participant diet records.

#### Phenol-Explorer database

As described previously^43^, Phenol-Explorer is a comprehensive online database describing (poly)phenol content of foods, collected from 637 peer-reviewed publications. The database includes content of 501 individual (poly)phenols in 458 plant-based food items. Phenol-Explorer assigns foods to 9 food groups including fruits, vegetables, cereal products, alcoholic beverages, coffee and cocoa products, seasonings, seeds, and oils. Individual (poly)phenols represent 5 classes (flavonoids, phenolic acids, lignans, stilbenes, and other (poly)phenols). The “other (poly)phenols” class includes other compounds such as alkylphenols, hydroxycoumarins, phenolic terpenes, and tyrosols. (Poly)phenols are also categorized into 14 subclasses: anthocyanins, dihydrochalcones, ellagitannins, flavan-3-ols, flavanones, flavones, flavonols, hydroxybenzoic acids, hydroxycinnamic acids, hydroxyphenylacetic acids, hydroxyphenylpropanoic acids, isoflavones, phenolic ketones, and proanthocyanidins.

(Poly)phenol content in Phenol-Explorer was determined using chromatographic methods (individual (poly)phenols), the Folin assay (total (poly)phenols), and the pH differential method (total anthocyanins).

#### Mapping between NDSR and Phenol-Explorer food items

Maternal and child diet records reported a total of 4,304 unique food items, with 2,229 food items identified as containing (poly)phenols. Examples of food items excluded due to having low (poly)phenol content include: salad dressings and sauces not derived primarily from plant-based foods, animal-based foods (e.g., eggs, poultry, beef, pork, fish and seafood, dairy), soft drinks, nutrient-enhanced water beverages, and candies that do not contain dark chocolate. Where possible, individual food items from NDSR were mapped directly to foods in the Phenol-Explorer database. Due to redundancy in food descriptions in Phenol-Explorer (e.g., 5 different types of blueberries), the database food items were manually assigned a food type (see **Table S1**) prior to mapping food items in NDSR. For cases in which items did not have a clear match in Phenol-Explorer, items were considered individually via 1 of 3 procedures that are fully documented in **Tables S1-S6**: substitution to assign a similar food item (e.g., Doritos, Sun Chips, and Tortilla chips were all assigned to the food type “Tortilla, white/yellow [chips, hard shell]”), generation of (poly)phenol profiles from published literature not represented in the Phenol-Explorer database (e.g., applesauce, honey, blackberry juice), or manually estimating (poly)phenol content using factors to adjust relevant data (e.g., using moisture content to calculate dried fruit (poly)phenol content from fresh fruit values). This resulted in supplementing the Phenol-Explorer data with an additional 36 references, 34 compounds, and 50 food types (accounts for 11% of individual food items and represents ∼40% of estimated total (poly)phenol intakes). For the remaining 320 food items found in NDSR, an appropriate match could not be found (e.g., potato chips, popcorn, snack mixes, plant-based alternative “milk” products, plant-based fats with uncertain composition). Thus, out of 2,229 NDSR food items identified as containing (poly)phenols, 1,909 food items (86%) were successfully mapped to an appropriate food type in the database.

#### Estimation of (poly)phenol intake

Total (poly)phenol content was determined as the sum of individual (poly)phenols measured using chromatographic methods. Class and subclass intakes were determined as the sum of individual (poly)phenols in each class or subclass, respectively (see **Table S2**). (Poly)phenol content determined using the Folin assay or pH differential method was excluded due to the non-specific, semi-quantitative nature of these assays. Prior to calculating (poly)phenol intakes, (poly)phenol content was averaged based on food type (see above), followed by aggregating data to determine the total (poly)phenol content, total of each (poly)phenol class, and total of each (poly)phenol subclass. Following aggregation, (poly)phenol intakes were calculated for each participant as the sum of the product of the amount of food (g/day) and the (poly)phenol content (mg/g) for a given food item (performed for each day of recorded dietary intake). Because energy-adjusted (poly)phenol intakes (mg/1000 kcal/d) did not differ across trimesters (1^st^ trimester: 371 ± 203; 2^nd^ trimester: 396 ± 274; 3^rd^ trimester: 384 ± 275; p = 0.59), (poly)phenol intakes were averaged across pregnancy for maternal intakes prior to statistical analyses. Three-day averages of energy-adjusted (poly)phenol intakes were calculated for child intake at age 5 and 8 years. Significant differences in child intake between age 5 and 8 years were identified using Wilcoxon signed-rank test. Top food contributors of total (poly)phenol intakes and each class and subclass was determined for pregnancy and child intakes by accounting for both food intake and (poly)phenol content of the food (g food/day x mg (poly)phenol/g food).

### Child cognitive assessments at 8 years

Standardized assessments were used to evaluate child cognitive performance at age 5 and 8 years across multiple domains of psychological function, including academic skills (Wide Range Achievement Test, WRAT4)^44^, language development (Clinical Evaluation of Language Fundamentals (CELF-5)^45^, and intelligence (Wechsler Abbreviated Scale of Intelligence (WASI-II)^46^. Psychological assessments were administered by a licensed psychological examiner or a neuropsychology technician supervised by a psychologist and neuropsychologist and were not administered to children who were out of the required age ranges for the assessments at the 5-year visit (WRAT4, 5.0-5.2 years, N = 6; CELF-5, 5.0-5.5 years, N = 2) or 8-year visit (WRAT4 and WASI-II, 8.0-8.3 years, N = 10; CELF-5, 8.0-8.11 years, N = 4). Scores below 80 were considered below average. Details on the assessments are provided here:

#### Academic achievement assessment

The WRAT4 provides a measure of basic academic skills including reading, spelling, and math computation and is validated for use at age 5 to 94 years^44^. Raw scores were converted to age-based standard scores having a mean of 100 and a standard deviation of 15, with scores ranging from 55 to 145.

#### Language development assessment

The CELF-5 is designed to assess the presence of language developmental delay^47^ and was validated for for use at age 5 to 21 years^45^. Scores of the CELF-5 included for analysis were Core Language, Receptive Language, and Expressive Language, which assess general language ability, listening and auditory comprehension, and expressive language skills, respectively. Like the WRAT4, the CELF-5 uses a standard mean score of 100 and a standard deviation of 15.

#### Intelligence assessment

The WASI-II was used to derive scores for verbal comprehension (verbal IQ; based on Vocabulary and Similarities subtests), perceptual reasoning (perceptual reasoning IQ; based on Block Design and Matrix Reasoning subtests), and full-scale IQ^46^. It is validated for use at age 6 to 90 years^46^. Verbal IQ is a measure of word meanings and verbal abstract reasoning, while perceptual reasoning IQ measures visuospatial reasoning.

### Maternal anthropometry

Maternal anthropometrics were measured at enrollment for the parent cohort (early pregnancy). Maternal weight and height were measured with a standing digital scale (Tanita Corporation, Arlington Heights, IL) and a wall-mounted stadiometer (Perspective Enterprises, Portage, MI), respectively. Body mass index (BMI, kg/m^2^) was calculated as weight (kg) / [height (m)]^2^. Gestational weight gain (GWG) was calculated based on weight gain between the first study visit and 36 weeks of gestation. The National Academy of Sciences, Engineering, and Medicine (NASEM) guidelines for GWG based on BMI were used to categorize women as having below (less than the recommended weight gain for BMI category), within (within the recommended weight gain for BMI category), and above (exceeding the recommended weight gain for BMI category) weight gain, while adjusting for gestation weeks^48^.

### Self-reported variables

At the time of participant enrollment, participants self-reported sociodemographic information including maternal age, educational attainment, and self-identified race/ethnicity. At 2 weeks postpartum, mothers reported infant race, sex, birth weight, birth length, and delivery mode. Gestational age was calculated using the mother’s last menstrual period and the child’s date of birth. Breastfeeding duration was calculated using the date of breastfeeding cessation and the child’s date of birth. Maternal IQ was assessed using the Wechsler Abbreviated Scale of Intelligence (WASI)^46^.

### Statistical analysis

Study demographics, anthropometrics, birth characteristics, breastfeeding duration, maternal IQ, child psychological assessment scores, and maternal and child dietary intake data were described using frequencies (%) for categorical variables or means and standard deviations (SDs) for continuous variables. Histograms were used to visualize the distribution of subtest scores for each psychological assessment: WRAT4 (math, spelling, reading), CELF-5 (core language, expressive language, receptive language), and WASI-II (verbal IQ, perceptual reasoning IQ, full scale IQ). Statistical significance was determined using a two-sided α level of 0.05 and uncertainty in parameter estimates was presented using two-sided 95% confidence intervals.

#### Associations between (poly)phenol intakes during pregnancy and (poly)phenol intakes at age 5 and 8 years

Total (poly)phenol intakes and individual (poly)phenol classes and subclasses during pregnancy that were significantly associated with child (poly)phenol intakes at age 5 and 8 years were identified using unadjusted linear mixed effects mdoels. Each model used the same maternal and child (poly)phenol measure (e.g., association between flavone intake during pregnancy and child flavone intake at age 5 and 8 years).

#### Variable selection for identifying (poly)phenols during pregnancy associated with cognitive outcomes at age 5 and 8 years

Given the large number of (poly)phenol classes and subclasses to consider, elastic net-based variable selection was completed using the glmmLASSO R package^49^ for longitudinal outcomes (WRAT4, CELF-5) and the glmnet R package for cross-sectional outcomes (WASI-II)^50^ using average (poly)phenol intakes during pregnancy. Variable selection was limited to mutually exclusive (poly)phenol classes and subclasses contributing ≥ 1% of total prenatal (poly)phenol intake (anthocyanins, flavan-3-ols, flavanones, flavones, flavonols, hydroxybenzoic acids, hydroxycinnamic acids, proanthocyanidins, lignans, and other (poly)phenols). Breastfeeding duration and maternal IQ were included as covariates for variable selection. Variables selected were then used in the multivariable regression models described below.

#### Linear models for predicting cognitive outcomes at age 5 and 8 years

Linear mixed effects models were used to evaluate associations between individual variables obtained from elastic net-based variable selection and longitudinal psychological assessments administered at age 5 and 8 years (WRAT4, CELF-5). Linear regression models were constructed for the psychological assessments administered at age 8 years (WASI-II). Each of the selected (poly)phenols was considered individually in the models. Model 1 was adjusted for breastfeeding duration and maternal IQ. Model 2 was additionally adjusted for pre-pregnancy BMI, gestational age, and infant sex. Model 3 was additionally adjusted for child (poly)phenol intake at age 5 and 8 years using the same (poly)phenol measure as the prenatal (poly)phenol predictor. For example, models that included prenatal intake of the (poly)phenol subclass anthocyanins as the predictor would include child intake of anthocyanins at age 5 and 8 years as a covariate. Results are presented as beta coefficients, 95% confidence intervals (CI), and p-values. Beta coefficients represent the estimated change in the mean outcome. All analyses were preformed using R version 4.5.3 (R Foundation for Statistical Computing).

## Results

### Maternal and child characteristics

Demographics and clinical characteristics of the mother-child dyads included in the analyses (Analytical Sample; N = 111) and those excluded from the analyses (Full Cohort Only; N = 209) are shown in **Table 1**. The cohort included in the analyses consisted of mostly White women (89.2%) who were approximately 30 years of age. On average, maternal BMI was 25.5 ± 4.4 kg/m^2^. Gestational weight gain was 11.8 ± 3.7 kg, with most women falling above the NASEM guidelines (above: 52.3%; within: 33.3%; below: 14.4%). All infants were full term at birth, with a mean gestational age at delivery of 39.4 ± 1.2 weeks. On average, children were breastfed for 7.4 ± 6.4 months. Overall, the characteristics of the mother-child dyads included in the analyses were similar to mother-child dyads that were excluded, with the exception of modestly lower BMI (p = 0.088) and a greater proportion of vaginal births (p = 0.05) in the analytical sample (BMI: 25.5 ± 4.4 kg/m^2^; 71.2% vaginal births) compared to participants only in the full cohort (BMI: 26.4 ± 4.4 kg/m^2^; 58.8% vaginal births). On average, children were 5.1 ± 0.1 years (range: 4.9-5.9 years) at the 5-year visit and 8.1 ± 0.1 years (range: 8.0-8.7 years) at the 8-year visit.

**Table 1.** Baseline characteristics of participants in the analytical cohort and full cohort.

| Characteristic | Analytical Sample<br>(N = 111) | Full Cohort Only<br>(N = 209) | All Participants<br>(N = 320) | p-value<sup>a</sup> |
| --- | --- | --- | --- | --- |
| Maternal age, years (SD) | 30.3 (3.3) | 30.1 (3.9) | 30.2 (3.6) | 0.75 |
| Maternal BMI, kg/m<sup>2</sup> (SD) | 25.5 (4.4) | 26.4 (4.4) | 26.1 (4.4) | 0.088 |
| Maternal race, N (%) |  |  |  | 0.78 |
| White | 99 (89.2%) | 173 (85.6%) | 272 (86.9%) |  |
| African American and others | 12 (10.8%) | 29 (14.4%) | 41 (13.1%) |  |
| Maternal education, N (%) |  |  |  | 0.49 |
| No College Degree | 7 (6.3%) | 13 (6.4%) | 20 (6.4%) |  |
| Partial College (1+ years) | 21 (18.9%) | 53 (26.1%) | 74 (23.6%) |  |
| College Degree/Specialized Training | 52 (46.8%) | 78 (38.4%) | 130 (41.4%) |  |
| Graduate Training/Degree | 31 (27.9%) | 59 (29.1%) | 90 (28.7%) |  |
| Household income, N (%) |  |  |  | 0.30 |
| <\$60,000 | 24 (22.0%) | 53 (27.5%) | 77 (25.5%) | |
| >\$60,000 | 85 (78.0%) | 140 (72.5%) | 225 (74.5%) | |
| GWG, kg (SD) | 11.8 (3.7) | 11.9 (4.5) | 11.9 (4.1) | 0.84 |
| NASEM GWG recommendations, N (%) |  |  |  | 0.65 |
| Above | 58 (52.3%) | 65 (52.8%) | 123 (52.6%) |  |
| Within | 37 (33.3%) | 45 (36.6%) | 82 (35.0%) |  |
| Below | 16 (14.4%) | 13 (10.6%) | 29 (12.4%) |  |
| Birth weight, kg (SD) | 3.5 (0.5) | 3.5 (0.5) | 3.5 (0.5) | 0.93 |
| Birth length, cm (SD) | 51.2 (2.6) | 51.0 (2.6) | 51.1 (2.6) | 0.61 |
| Infant sex, N (%) |  |  |  | 0.30 |
| Female | 51 (45.9%) | 47 (39.2%) | 98 (42.4%) |  |
| Male | 60 (54.1%) | 73 (60.8%) | 133 (57.6%) |  |
| Gestational age, weeks (SD) | 39.4 (1.2) | 39.3 (1.0) | 39.3 (1.1) | 0.83 |
| Delivery method, N (%) |  |  |  | 0.050 |
| Vaginal | 79 (71.2%) | 70 (58.8%) | 149 (64.8%) |  |
| C-section | 32 (28.8%) | 49 (41.2%) | 81 (35.2%) |  |
| Breastfeeding duration, months (IQR) | 6.2 (11.1) | 5.5 (12.3) | 5.9 (11.5) | 0.76 |
| Maternal full-scale IQ (SD) | 107.6 (10.6) | 106.4 (9.5) | 107.0 (10.1) | 0.36 |

Data presented as counts (%) or means (SD). Breastfeeding duration is presented as medians (IQR) because it is not normally distributed.
<sup>a</sup>Differences between the analytical sample and participants only in the full cohort were tested using a t-test for continuous variables and a chi square test for categorical variables. Differences for breastfeeding duration were tested using the Mann-Whitney U test.
GWG, gestational weight gain

### (Poly)phenol intakes during pregnancy and childhood

(Poly)phenol intakes averaged across pregnancy and major food contributors to intakes are shown in **Table 2**. Unadjusted mean total (poly)phenol intake during pregnancy was 716 ± 383 mg/d. The most consumed (poly)phenol classes were flavonoids (53% of total intake) and phenolic acis (42% of total intake). The highest consumed subclasses were hydroxycinnamic acids, comprising 38% of all (poly)phenols with a majority of intake contributed by coffee, corn bran, and orange juice. Flavan-3-ols were the second highest consumed during pregnancy with beans, blueberries, and green tea being the major dietary contributors. Flavonols contributed 16% to total intake, primarily from coffee, orange juice, and herbal tea intake, while proanthocyanidins comprised 10% of total intake, mostly derived from beans, plums, and dark chocolate.

**Table 2.**
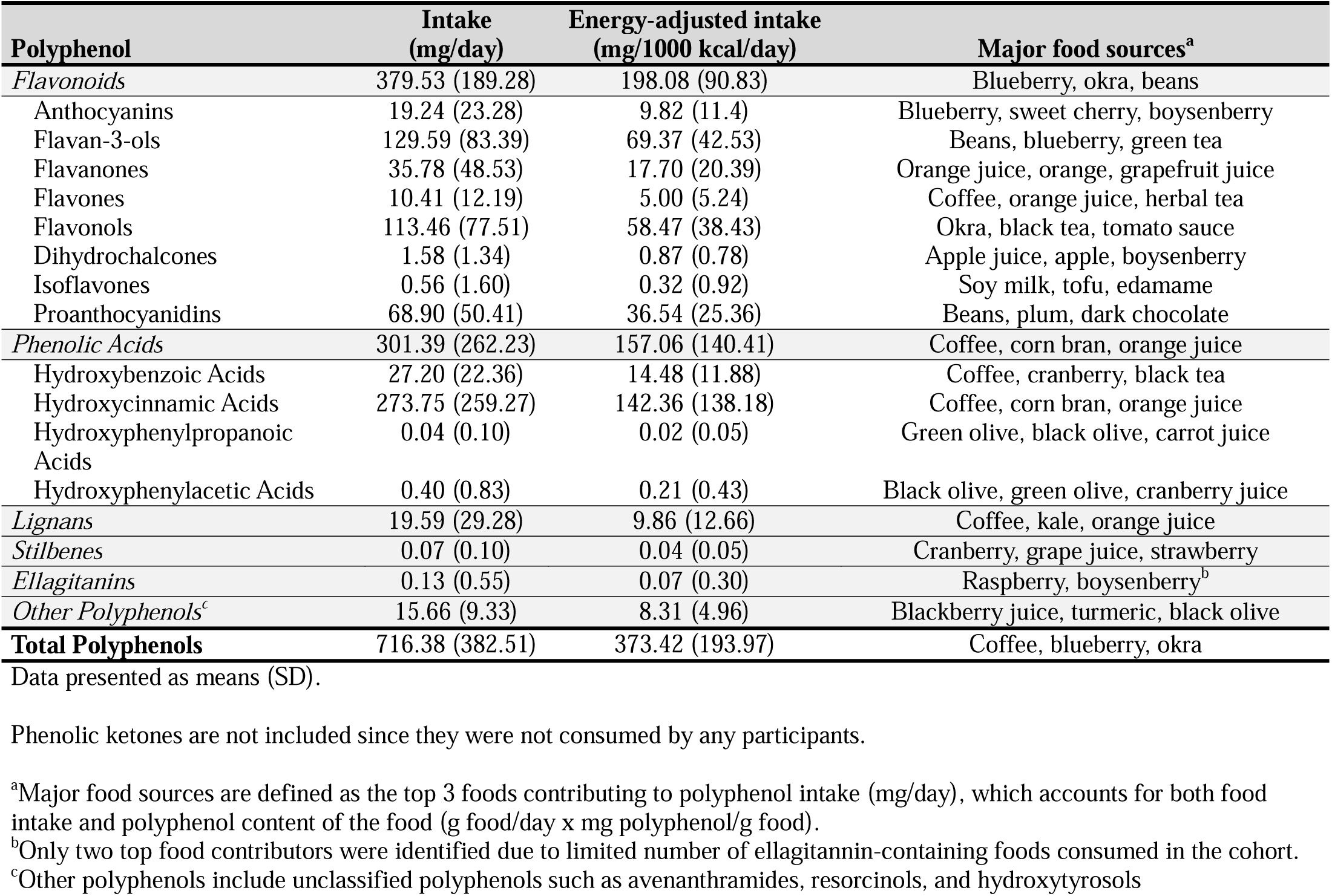
Intakes (mg/day), energy-adjusted intakes (mg/1000 kcal/day), and major food sources of polyphenols consumed during pregnancy.

| **Polyphenol** | **Intake**<br>**(mg/day)** | **Energy-adjusted intake**<br>**(mg/1000 kcal/day)** | **Major food sources<sup>a</sup>** |
| --- | --- | --- | --- |
| *Flavonoids* | 379.53 (189.28) | 198.08 (90.83) | Blueberry, okra, beans |
| Anthocyanins | 19.24 (23.28) | 9.82 (11.4) | Blueberry, sweet cherry, boysenberry |
| Flavan-3-ols | 129.59 (83.39) | 69.37 (42.53) | Beans, blueberry, green tea |
| Flavanones | 35.78 (48.53) | 17.70 (20.39) | Orange juice, orange, grapefruit juice |
| Flavones | 10.41 (12.19) | 5.00 (5.24) | Coffee, orange juice, herbal tea |
| Flavonols | 113.46 (77.51) | 58.47 (38.43) | Okra, black tea, tomato sauce |
| Dihydrochalcones | 1.58 (1.34) | 0.87 (0.78) | Apple juice, apple, boysenberry |
| Isoflavones | 0.56 (1.60) | 0.32 (0.92) | Soy milk, tofu, edamame |
| Proanthocyanidins | 68.90 (50.41) | 36.54 (25.36) | Beans, plum, dark chocolate |
| *Phenolic Acids* | 301.39 (262.23) | 157.06 (140.41) | Coffee, corn bran, orange juice |
| Hydroxybenzoic Acids | 27.20 (22.36) | 14.48 (11.88) | Coffee, cranberry, black tea |
| Hydroxycinnamic Acids | 273.75 (259.27) | 142.36 (138.18) | Coffee, corn bran, orange juice |
| Hydroxyphenylpropanoic Acids | 0.04 (0.10) | 0.02 (0.05) | Green olive, black olive, carrot juice |
| Hydroxyphenylacetic Acids | 0.40 (0.83) | 0.21 (0.43) | Black olive, green olive, cranberry juice |
| *Lignans* | 19.59 (29.28) | 9.86 (12.66) | Coffee, kale, orange juice |
| *Stilbenes* | 0.07 (0.10) | 0.04 (0.05) | Cranberry, grape juice, strawberry |
| *Ellagitannins* | 0.13 (0.55) | 0.07 (0.30) | Raspberry, boysenberry<sup>b</sup> |
| *Other Polyphenols<sup>c</sup>* | 15.66 (9.33) | 8.31 (4.96) | Blackberry juice, turmeric, black olive |
| **Total Polyphenols** | **716.38 (382.51)** | **373.42 (193.97)** | Coffee, blueberry, okra |

Data presented as means (SD).
Phenolic ketones are not included since they were not consumed by any participants.
<sup>a</sup>Major food sources are defined as the top 3 foods contributing to polyphenol intake (mg/day), which accounts for both food intake and polyphenol content of the food (g food/day x mg polyphenol/g food).
<sup>b</sup>Only two top food contributors were identified due to limited number of ellagitannin-containing foods consumed in the cohort.
<sup>c</sup>Other polyphenols include unclassified polyphenols such as avenanthramides, resorcinols, and hydroxytyrosols

(Poly)phenol intakes in children at age 5 and 8 years and major food contributors to intakes are shown in **Table 3** and **Table 4**. Unadjusted mean total (poly)phenol intake at age 5 was 395 ± 221 mg/d and at age 8 was 411 ± 262 mg/d. Qualitatively, profiles of (poly)phenol intake in children were similar to maternal intakes during pregnancy. At age 5 and 8 years, the (poly)phenol classes contributing the most to total intake were flavonoids (5 years: 66%; 8 years: 69%) and phenolic acids (5 years: 29%; 8 years: 27%). Hydroxycinnamic acids (5 years: 26%; 8 years: 23%) and flavan-3-ols (5 years: 23%; 8 years: 27%) were consumed the most, followed by flavonols (5 years: 16%; 8 years: 14%) and proanthocyanidins (5 years: 13%; 8 years: 16%). Certain contributors to the majority of (poly)phenol intakes in children were consistent at both 5 and 8 years of age, while others varied. At both 5 and 8 years of age, orange juice and plums were the primary source of hydroxycinnamic acids, while beans were the major contributor of flavan-3-ols. Most proanthocyanidins were consumed through intake of beans, plums, and dark chocolate. Flavonols were primarily provided through intake of okra and tomato sauce. Major food sources that varied between age 5 and 8 included plums as a major contributor of flavan-3-ols at age 5, while blueberries were a primary food source at age 8. Beans were a major contributor to flavonol intakes at age 5, but black tea (e.g., sweetened or unsweetened black tea) was a major contributor at age 8. Fresh sweet cherry was a major source of hydroxycinnamic acids at age 5, while blueberries were a primary contributor at age 8. Intakes of most (poly)phenols did not differ between age 5 and 8, with the exception of energy-adjusted phenolic acid, hydroxycinnamic acid, and flavanone intakes, and unadjusted flavan-3-ol intakes. Total phenolic acid intakes were lower at age 8 (63 ± 28 mg/1000 kcal/d) compared to age 5 (77 ± 33 mg/1000 kcal/d; p = 0.004). This difference in intake was driven by hydroxycinnamic acid intakes, which were lower at age 8 (54 ± 25 mg/1000 kcal/d) compared to age 5 (68 ± 31 mg/1000 kcal/d; p = 0.004). Flavanone intakes were also lower at age 8 (11 ± 18 mg/1000 kcal/d) compared to age 5 (21 ± 27 mg/1000 kcal/d; p = 0.004), while flavan-3-ol intakes were higher at age 8 (109 ± 94 mg/d) compared to age 5 (89 ± 81 mg/d; p = 0.008).

**Table 3.** Intakes (mg/day), energy-adjusted intakes (mg/1000 kcal/day), and major food sources of polyphenols consumed by children at age 5 years of age (N = 78)

| **Polyphenol** | **mg/day** | **mg/1000 kcal/day** | **Major food sources<sup>a</sup>** |
| --- | --- | --- | --- |
| *Flavonoids* | 261.78 (183.10) | 178.85 (132.73) | Okra, beans, plum |
| Anthocyanins | 20.01 (28.85) | 13.72 (19.28) | Sweet cherry, blackberry, blueberry |
| Flavan-3-ols | 89.08 (80.77)* | 60.75 (56.06) | Beans, plum, apple |
| Flavanones | 31.23 (41.88) | 20.67 (26.94)* | Orange juice, grapefruit, orange juice |
| Flavones | 5.50 (4.34) | 3.58 (2.72) | Parsley, orange juice, black olive |
| Flavonols | 62.51 (86.55) | 43.29 (64.57) | Okra, tomato sauce, beans |
| Dihydrochalcones | 3.58 (5.16) | 2.32 (3.21) | Apple juice, apple, applesauce |
| Isoflavones | 0.11 (0.39) | 0.08 (0.26) | Soy flour, edamame, beans |
| Proanthocyanidins | 49.76 (47.16) | 34.45 (34.91) | Beans, plum, dark chocolate |
| *Phenolic Acids* | 115.22 (49.96) | 77.04 (33.28)* | Coffee, orange juice, plum |
| Hydroxybenzoic Acids | 12.96 (9.92) | 8.98 (7.34) | Okra, black olive, blackberry |
| Hydroxycinnamic Acids | 101.98 (48.20) | 67.84 (31.28)* | Orange juice, plum, sweet cherry |
| Hydroxyphenylpropanoic Acids | 0.02 (0.12) | 0.02 (0.09) | Black olive, vinegar<sup>b</sup> |
| Hydroxyphenylacetic Acids | 0.26 (1.26) | 0.19 (0.91) | Black olive, cranberry juice, RG corn flour |
| *Lignans* | 10.35 (11.25) | 7.32 (8.08) | Broccoli, kale, orange juice |
| *Stilbenes* | 0.06 (0.09) | 0.04 (0.06) | Strawberry, grape, cranberry |
| *Ellagitannins* | 0.23 (1.04) | 0.15 (0.68) | Raspberry<sup>b</sup> |
| *Other Polyphenols<sup>c</sup>* | 7.64 (9.24) | 5.25 (6.99) | Black olive, WG pasta, cloves |
| **Total Polyphenols** | **395.28 (220.82)** | **286.65 (158.47)** | **Okra, beans, plum** |

Data presented as means (SD).
Phenolic ketones are not included since they were not consumed by any participants.
\*Significantly different from intake at age 8 years ( $p < 0.05$ ).
<sup>a</sup>Major food sources are defined as the top 3 foods contributing to polyphenol intake (mg/day), which accounts for both food intake and polyphenol content of the food (g food/day x mg polyphenol/g food)
<sup>b</sup><3 top food contributors were identified due to limited number of foods containing the polyphenol (e.g., ellagitannins) consumed in the cohort.
<sup>c</sup>Other polyphenols include unclassified polyphenols such as avenanthramides, resorcinols, and hydroxytyrosols
WG, whole grain; RG, refined grain

**Table 4.** Intakes (mg/day), energy-adjusted intakes (mg/1000 kcal/day), and major food sources of polyphenols consumed by children at age 8 years of age (N = 80)

| **Polyphenol** | **mg/day** | **mg/1000 kcal/day** | **Major food sources<sup>a</sup>** |
| --- | --- | --- | --- |
| *Flavonoids* | 282.28 (218.28) | 160.77 (124.80) | Beans, okra, green tea |
| Anthocyanins | 21.74 (31.88) | 12.82 (20.29) | Blueberry, blackberry, strawberry |
| Flavan-3-ols | 109.26 (94.33) | 63.04 (54.44) | Beans, blueberry, apple |
| Flavanones | 20.46 (32.36) | 11.18 (17.55) | Orange juice, orange, lemon juice |
| Flavones | 6.03 (5.15) | 3.33 (2.78) | Orange juice, WG wheat flour, parsley |
| Flavonols | 58.45 (56.37) | 31.73 (28.50) | Okra, black tea, tomato sauce |
| Dihydrochalcones | 2.50 (3.45) | 1.47 (2.15) | Apple, apple juice, applesauce |
| Isoflavones | 0.13 (0.46) | 0.08 (0.33) | Tofu, soy flour, chia |
| Proanthocyanidins | 63.70 (64.42) | 37.12 (37.34) | Beans, dark chocolate, plum |
| *Phenolic Acids* | 111.01 (48.97) | 62.81 (28.15) | Orange juice, blueberry, plum |
| Hydroxybenzoic Acids | 15.30 (16.79) | 8.59 (8.88) | Black tea, okra, strawberry |
| Hydroxycinnamic Acids | 95.57 (42.44) | 54.13 (24.78) | Orange juice, blueberry, plum |
| Hydroxyphenylpropanoic Acids | 0.01 (0.05) | 0.01 (0.04) | Black olive, vinegar<sup>b</sup> |
| Hydroxyphenylacetic Acids | 0.12 (0.57) | 0.08 (0.39) | Black olive, RG corn flour, olive oil |
| *Lignans* | 10.92 (11.10) | 6.32 (6.48) | Orange juice, sesame oil, peach |
| *Stilbenes* | 0.08 (0.14) | 0.05 (0.07) | Strawberry, grape, peanut |
| *Ellagitannins* | 0.07 (0.43) | 0.04 (0.23) | Raspberry<sup>b</sup> |
| *Other Polyphenols<sup>c</sup>* | 6.56 (6.84) | 3.77 (4.07) | Black olive, WG pasta, WG rye bread |
| **Total Polyphenols** | **410.93 (261.75)** | **233.76 (148.63)** | **Beans, orange juice, blueberry** |

Data presented as means (SD).
Phenolic ketones are not included since they were not consumed by any participants.
<sup>a</sup>Major food sources are defined as the top 3 foods contributing to polyphenol intake (mg/day), which accounts for both food intake and polyphenol content of the food (g food/day x mg polyphenol/g food)
<sup>b</sup><3 top food contributors were identified due to limited number of foods containing the polyphenol (e.g., ellagitannins) consumed in the cohort.
<sup>c</sup>Other polyphenols include unclassified polyphenols such as avenanthramides, resorcinols, and hydroxytyrosols
WG, whole grain; RG, refined grain

### Higher prenatal (poly)phenol intake associates positively with (poly)phenol intakes in children

Relationships between prenatal (poly)phenol intakes and (poly)phenol intakes in children at 5 and 8 years of age (total, class, and subclass-level) were tested using linear mixed effect models that included log-transformed and energy-adjusted prenatal and child intakes of the same (poly)phenol measure (e.g., association between prenatal flavone intake and child flavone intake). Outputs from all of the models are presented in **Table S6**.

Linear mixed effect models revealed that higher prenatal (poly)phenol intakes were associated with higher child (poly)phenol intakes for lignans (p = 0.007), other (poly)phenols (p = 0.007; uncategorized phenolics including avenanthramides, catechol, coumestrol, curcumin, and tyrosol), and proanthocyanidins (p = 0.04). Increasing prenatal intakes of proanthocyanidins, lignans, and other (poly)phenols by 10% corresponded to a 3% increase in child intakes of these (poly)phenols.

### Academic and neurocognitive performance of children at age 5 and 8 years

The academic and neurocognitive test scores of the children at age 5 and 8 years are shown in **Table 5** and **Figure 3**. Overall, results indicated that children’s scores on all administered assessments were at or above average (standard scores ≥ 80)^44–46^. Performance for the majority of this cohort tended to fall at or above average ranges on the academic and neurocognitive measures. For most assessments, over 90% of children scored at or above average ranges. The lowest scores were observed for reading skills at age 5, with 17.2% of children scoring below average. However, reading scores improved by 8 years of age, with only 2.7% of children scoring below average. The assessments with the least proportion of children scoring below average were core language scores at age 5 (1.1%) and verbal IQ scores at age 8 (0%).

**Figure 3:**
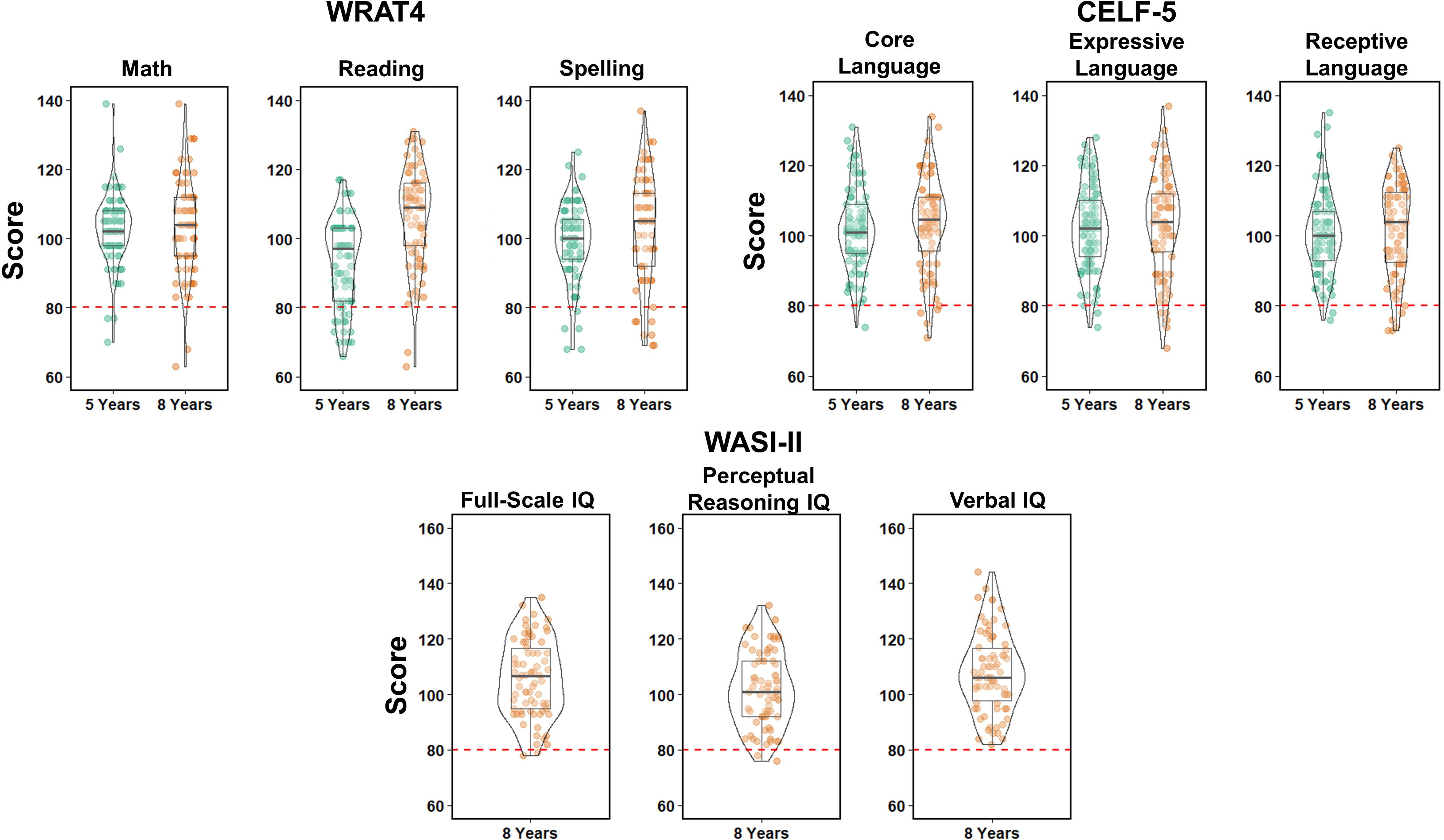
Violin plots showing distributions for *T*-scores from the Wide Range Achievement Test, 4^th^ edition^43^ (WRAT4; math, reading, and spelling) at age 5 and 8 years, Clinical Evaluation of Language Fundamentals, 5^th^ edition^44^ (CELF-5; core, expressive, and receptive language) at age 5 and 8 years, and Wechsler Abbreviatedd Scale of Intelligence, 2^nd^ edition^45^ (WASI-II; full-scale, perceptual reasoning, and verbal IQ). The red dashed line indicates cutoff for scores that are “Below Average” (< 80).

**Table 5.** Academic and neurocognitive performance of children at age 5 and 8 years.

| Assessment | 5 years |  |  | 8 years |  |  |
| --- | --- | --- | --- | --- | --- | --- |
|  | *Below*<br>*Average (%)* | *At or Above*<br>*Average (%)* | *Mean (SD)* | *Below*<br>*Average (%)* | *At or Above*<br>*Average (%)* | *Mean (SD)* |
| **WRAT4** |  |  |  |  |  |  |
| Reading score | 15 (17.2%) | 72 (82.8%) | 93.2 (12.4) | 2 (2.7%) | 72 (97.3%) | 106.5 (14.3) |
| Spelling score | 5 (5.7%) | 82 (94.3%) | 99.1 (11.0) | 7 (9.9%) | 64 (90.1%) | 103.2 (15.7) |
| Math score | 3 (3.4%) | 84 (96.6%) | 102.8 (9.8) | 2 (2.7%) | 72 (97.3%) | 103.3 (13.6) |
| **CELF-5** |  |  |  |  |  |  |
| Core language score | 1 (1.1%) | 86 (98.9%) | 101.8 (11.7) | 4 (5.3%) | 72 (94.7%) | 103.6 (13.2) |
| Expressive language score | 2 (2.3%) | 85 (97.7%) | 102.0 (12.0) | 5 (6.6%) | 71 (93.4%) | 103.2 (14.3) |
| Receptive language score | 2 (2.3%) | 85 (97.7%) | 100.5 (11.6) | 5 (6.4%) | 73 (93.6%) | 101.6 (13.0) |
| **WASI-II** |  |  |  |  |  |  |
| Full-scale IQ | --- | --- | --- | 2 (2.7%) | 72 (97.3%) | 105.8 (13.8) |
| Perceptual reasoning IQ | --- | --- | --- | 2 (2.7%) | 72 (97.3%) | 101.9 (13.3) |
| Verbal IQ | --- | --- | --- | 0 (0%) | 74 (100%) | 108.0 (14.3) |

Data presented as counts (%).
WRAT4, Wide Range Achievement Test, 4<sup>th</sup> edition<sup>43</sup>
CELF-5, Clinical Evaluation of Language Fundamentals, 5<sup>th</sup> edition<sup>44</sup>
WASI-II, Wechsler Abbreviated Scale of Intelligence, 2<sup>nd</sup> edition<sup>45</sup>

### Variable selection for associations of prenatal (poly)phenol intakes with child cognitive assessment scores

To identify the most important prenatal (poly)phenol predictors of child cognitive outcomes at age 5 and 8 years, elastic net-based variable selection was performed. The selected variables for each cognitive outcome are shown in **Table S7**. Each of the 10 (poly)phenols were selected for academic achievement scores (WRAT4). Each of the (poly)phenols except flavanones were selected for language scores (CELF-5). Flavones, lignans, and hydroxycinnamic acids were selected for IQ scores (WASI-II).

### Multivariable regression analysis of associations between prenatal (poly)phenol intakes and child cognitive function

The associations between the selected (poly)phenol predictors and each cognitive outcome were then further examined using multivariable regression. Results from linear mixed effect models associating prenatal (poly)phenol intakes with academic achievement scores (reading, spelling, and math) at 5 and 8 years of age from the WRAT4 assessment are shown in **Table 6**. In the fully adjusted models, there was a trend toward significance for positive associations of prenatal flavan-3-ol intakes with reading (β: 3.39 [−0.05, 6.82], p = 0.053) and spelling scores (β: 3.26 [−0.44, 6.95], p = 0.083). There was also a trend toward significance for positive associations of prenatal proanthocyanidin intakes with spelling scores (β: 3.14 [−0.12, 6.40], p = 0.059). Prenatal (poly)phenol intakes were not significantly associated with math scores.

**Table 6.** Linear mixed effect models for associations between selected prenatal polyphenol predictors and academic achievement outcomes in children at age 5 and 8 years.

| **Polyphenol** | **$\beta$** | **95% CI** | **p-value** |
| --- | --- | --- | --- |
| *Reading* |  |  |  |
| *Anthocyanins* |  |  |  |
| Model 1 | 1.010 | [-1.342, 3.361] | 0.40 |
| Model 2 | 1.125 | [-1.251, 3.502] | 0.35 |
| Model 3 | 1.030 | [-1.525, 3.585] | 0.43 |
| *Flavan-3-ols* |  |  |  |
| Model 1 | 3.220 | [0.090, 6.349] | 0.044 |
| Model 2 | 3.179 | [-0.018, 6.377] | 0.051 |
| Model 3 | 3.387 | [-0.045, 6.819] | 0.053 |
| *Flavanones* |  |  |  |
| Model 1 | -0.553 | [-3.049, 1.943] | 0.66 |
| Model 2 | -0.563 | [-3.079, 1.953] | 0.66 |
| Model 3 | -0.414 | [-3.164, 2.335] | 0.77 |
| *Flavonols* |  |  |  |
| Model 1 | 0.997 | [-2.630, 4.625] | 0.59 |
| Model 2 | 0.597 | [-3.176, 4.370] | 0.75 |
| Model 3 | 1.141 | [-2.979, 5.261] | 0.58 |
| *Proanthocyanidins* |  |  |  |
| Model 1 | 1.843 | [-0.952, 4.637] | 0.19 |
| Model 2 | 1.852 | [-0.965, 4.669] | 0.20 |
| Model 3 | 2.218 | [-0.851, 5.286] | 0.16 |
| *Hydroxybenzoic Acids* |  |  |  |
| Model 1 | 1.464 | [-1.835, 4.762] | 0.38 |
| Model 2 | 1.333 | [-2.026, 4.691] | 0.43 |
| Model 3 | 0.894 | [-2.679, 4.467] | 0.62 |
| *Hydroxycinnamic Acids* |  |  |  |
| Model 1 | 0.158 | [-2.889, 3.205] | >0.9 |
| Model 2 | 0.079 | [-2.998, 3.156] | >0.9 |
| Model 3 | -0.294 | [-3.659, 3.071] | 0.86 |
| *Lignans* |  |  |  |
| Model 1 | -0.416 | [-3.699, 2.867] | 0.80 |
| Model 2 | -0.296 | [-3.619, 3.027] | 0.86 |
| Model 3 | 0.598 | [-2.899, 4.096] | 0.74 |
| *Other Polyphenols* |  |  |  |
| Model 1 | 2.122 | [-2.090, 6.335] | 0.32 |
| Model 2 | 2.102 | [-2.163, 6.367] | 0.33 |
| Model 3 | 2.603 | [-2.143, 7.348] | 0.28 |
| *Spelling* |  |  |  |
| *Flavan-3-ols* |  |  |  |
| Model 1 | 2.825 | [-0.475, 6.125] | 0.092 |
| Model 2 | 3.076 | [-0.264, 6.417] | 0.071 |
| Model 3 | 3.255 | [-0.435, 6.946] | 0.083 |
| *Flavanones* |  |  |  |
| Model 1 | -1.265 | [-3.885, 1.355] | 0.34 |
| Model 2 | -1.344 | [-3.970, 1.282] | 0.31 |
| Model 3 | -1.260 | [-4.240, 1.720] | 0.40 |
| *Proanthocyanidins* |  |  |  |
| Model 1 | 2.864 | [-0.052, 5.781] | 0.054 |
| Model 2 | 2.822 | [-0.100, 5.745] | 0.058 |
| Model 3 | 3.140 | [-0.117, 6.398] | 0.059 |
| *Other Polyphenols* |  |  |  |
| Model 1 | 0.748 | [-3.704, 5.200] | 0.74 |
| Model 2 | 0.609 | [-3.878, 5.096] | 0.79 |
| Model 3 | 0.855 | [-4.365, 6.075] | 0.75 |
| *Math* |  |  |  |
| *Flavones* |  |  |  |
| Model 1 | 2.068 | [-1.460, 5.595] | 0.25 |
| Model 2 | 2.273 | [-1.273, 5.820] | 0.21 |
| Model 3 | 3.268 | [-0.814, 7.351] | 0.12 |
| *Other Polyphenols* |  |  |  |
| Model 1 | 0.708 | [-3.110, 4.527] | 0.71 |
| Model 2 | 1.008 | [-2.848, 4.865] | 0.61 |
| Model 3 | 1.298 | [-3.211, 5.807] | 0.57 |

Estimates represent the association between prenatal polyphenol intake (averaged across pregnancy) and child cognitive outcomes (age 5 and 8 years) using linear mixed effect models. Polyphenol intakes are energy-adjusted and log-transformed.
Model 1: Adjusted for maternal IQ and breastfeeding duration.
Model 2: Adjusted for the covariates in Model 2 plus maternal pre-pregnancy BMI, gestational age, and infant sex.
Model 3: Adjusted for the covariates in Model 2 plus child polyphenol intake using the same polyphenol measure as the prenatal polyphenol predictor.
Model 1 and 2 with reading or math scores as the outcome, N = 108; Model 1 and 2 with spelling scores as the outcome, N = 107; Model 3 with reading or math scores as the outcome, N = 100; Model 3 with spelling scores as the outcome, N = 99.

Results from linear mixed effect models used to evaluate associations between prenatal (poly)phenol intakes and language skills (core, receptive, and expressive) at 5 and 8 years of age are summarized in **Table 7**. Higher prenatal flavone intakes significantly associated with increased core (β: 4.36 [0.09, 8.62], p = 0.045) and expressive language scores (β: 4.55 [0.09, 9.02], p = 0.046). Higher prenatal flavonol intakes significantly associated with decreased core (β: −4.67 [−8.64, −0.70], p = 0.022) and expressive language scores (β: −4.24 [−8.41, −0.08], p = 0.046). Higher prenatal hydroxybenzoic acids intakes significantly associated with decreased core language scores (β: −3.88 [−7.29, −0.46], p = 0.026). A trend toward significance was observed for negative associations between prenatal hydroxybenzoic acid intakes and expressive language scores (β: −3.31 [−6.91, 0.30], p = 0.071). Prenatal (poly)phenol intakes were not significantly associated with receptive language scores.

**Table 7.** Linear mixed effect models for associations between selected prenatal polyphenol predictors and language skills outcomes in children at age 5 and 8 years.

| **Polyphenol** | **$\beta$** | **95% CI** | **p-value** |
| --- | --- | --- | --- |
| *Core Language* |  |  |  |
| *Flavones* |  |  |  |
| **Model 1** | **3.958** | **[0.149, 7.766]** | **0.042** |
| **Model 2** | **3.737** | **[-0.099, 7.574]** | **0.056** |
| **Model 3** | **4.355** | **[0.089, 8.621]** | **0.045** |
| *Flavonols* |  |  |  |
| **Model 1** | **-3.381** | **[-6.394, 0.172]** | **0.062** |
| **Model 2** | **-3.782** | **[-7.439, -0.126]** | **0.043** |
| **Model 3** | **-4.671** | **[-8.640, -0.701]** | **0.022** |
| *Proanthocyanidins* |  |  |  |
| Model 1 | 1.255 | [-1.552, 4.062] | 0.38 |
| Model 2 | 1.199 | [-1.618, 4.017] | 0.40 |
| Model 3 | 1.197 | [-1.837, 4.231] | 0.44 |
| *Hydroxybenzoic Acids* |  |  |  |
| **Model 1** | **-3.124** | **[-6.319, 0.072]** | **0.055** |
| **Model 2** | **-3.001** | **[-6.244, 0.242]** | **0.069** |
| **Model 3** | **-3.876** | **[-7.289, -0.464]** | **0.026** |
| *Hydroxycinnamic Acids* |  |  |  |
| Model 1 | 1.288 | [-1.728, 4.303] | 0.40 |
| Model 2 | 1.145 | [-1.890, 4.180] | 0.46 |
| Model 3 | 0.742 | [-2.611, 4.094] | 0.66 |
| *Lignans* |  |  |  |
| Model 1 | 0.813 | [-2.399, 4.029] | 0.62 |
| Model 2 | 0.667 | [-2.565, 3.899] | 0.68 |
| Model 3 | 0.634 | [-2.813, 4.080] | 0.72 |
| *Other Polyphenols* |  |  |  |
| Model 1 | 3.174 | [-0.949, 7.298] | 0.13 |
| Model 2 | 2.863 | [-1.316, 7.042] | 0.18 |
| Model 3 | 3.662 | [-0.983, 8.307] | 0.12 |
| *Receptive Language* |  |  |  |
| *Anthocyanins* |  |  |  |
| Model 1 | -0.097 | [-2.413, 2.219] | >0.9 |
| Model 2 | -0.052 | [-2.399, 2.295] | >0.9 |
| Model 3 | -0.011 | [-2.590, 2.568] | >0.9 |
| *Flavan-3-ols* |  |  |  |
| Model 1 | 0.150 | [-3.028, 3.327] | >0.9 |
| Model 2 | 0.277 | [-2.975, 3.528] | 0.87 |
| Model 3 | 0.141 | [-3.408, 3.690] | >0.9 |
| *Proanthocyanidins* |  |  |  |
| Model 1 | 1.683 | [-1.108, 4.474] | 0.23 |
| Model 2 | 1.691 | [-1.130, 4.513] | 0.24 |
| Model 3 | 1.793 | [-1.277, 4.863] | 0.25 |
| *Lignans* |  |  |  |
| Model 1 | 0.688 | [-2.517, 3.894] | 0.67 |
| Model 2 | 0.752 | [-2.498, 4.001] | 0.65 |
| Model 3 | 0.741 | [-2.766, 4.249] | 0.68 |
| *Expressive Language* |  |  |  |
| *Flavones* |  |  |  |
| **Model 1** | **3.857** | **[-0.194, 7.909]** | **0.062** |
| **Model 2** | **3.591** | **[-0.481, 7.663]** | **0.083** |
| **Model 3** | **4.550** | **[0.085, 9.015]** | **0.046** |
| *Flavonols* |  |  |  |
| **Model 1** | **-3.206** | **[-6.988, 0.576]** | **0.096** |
| **Model 2** | **-3.603** | **[-7.486, 0.281]** | **0.069** |
| **Model 3** | **-4.243** | **[-8.409, -0.076]** | **0.046** |
| *Proanthocyanidins* |  |  |  |
| Model 1 | 0.636 | [-2.348, 3.620] | 0.67 |
| Model 2 | 0.572 | [-2.415, 3.559] | 0.71 |
| Model 3 | 0.477 | [-2.698, 3.651] | 0.77 |
| *Hydroxybenzoic Acids* |  |  |  |
| Model 1 | -2.578 | [-5.990, 0.835] | 0.14 |
| Model 2 | -2.405 | [-5.859, 1.049] | 0.17 |
| Model 3 | -3.310 | [-6.914, 0.295] | 0.071 |
| *Hydroxycinnamic Acids* |  |  |  |
| Model 1 | 1.791 | [-1.400, 4.983] | 0.27 |
| Model 2 | 1.633 | [-1.572, 4.838] | 0.32 |
| Model 3 | 0.888 | [-2.611, 4.387] | 0.62 |
| *Lignans* |  |  |  |
| Model 1 | 0.634 | [-2.775, 4.042] | 0.71 |
| Model 2 | 0.453 | [-2.968, 3.873] | 0.79 |
| Model 3 | 0.382 | [-3.219, 3.982] | 0.83 |
| *Other Polyphenols* |  |  |  |
| Model 1 | 3.281 | [-1.096, 7.657] | 0.14 |
| Model 2 | 2.907 | [-1.518, 7.331] | 0.20 |
| Model 3 | 3.332 | [-1.515, 8.179] | 0.18 |

Estimates represent the association between prenatal polyphenol intake (averaged across pregnancy) and child cognitive outcomes (age 5 and 8 years) using linear mixed effect models. Polyphenol intakes are energy-adjusted and log-transformed.
Model 1: Adjusted for maternal IQ and breastfeeding duration.
Model 2: Adjusted for the covariates in Model 2 plus maternal pre-pregnancy BMI, gestational age, and infant sex.
Model 3: Adjusted for the covariates in Model 2 plus child polyphenol intake using the same polyphenol measure as the prenatal polyphenol predictor.
Model 1 and 2, N = 108; Model 3, N = 102.

Linear regression models used to evaluate associations between prenatal (poly)phenol intakes and IQ scores at 8 years of age using the WASI-II assessment are shown in **Table 8**. In the fully adjusted models, higher prenatal intakes of flavones significantly associated with increased full-scale (β: 9.43 [3.50, 15.36], p = 0.002), perceptual reasoning (β: 6.35 [0.22, 12.48], p = 0.042), and verbal IQ scores (β: 10.24 [3.87, 16.61], p = 0.002). Higher prenatal lignan intakes, derived primarily from coffee, kale, and orange juice, were significantly associated with increased verbal IQ scores (β: 10.24 [3.87, 13.61], p = 0.002).

**Table 8.** Linear regression models for associations between selected prenatal polyphenol predictors and IQ score outcomes in children at age 8 years.

| **Polyphenol** | **$\beta$** | **95% CI** | ***p*-value** |
| --- | --- | --- | --- |
| *Full Scale IQ* |  |  |  |
| *Flavones* |  |  |  |
| **Model 1** | **9.384** | **[4.183, 14.585]** | **<0.001** |
| **Model 2** | **9.421** | **[4.090, 14.753]** | **<0.001** |
| **Model 3** | **9.428** | **[3.500, 15.355]** | **0.002** |
| *Perceptual Reasoning IQ* |  |  |  |
| *Flavones* |  |  |  |
| **Model 1** | **7.310** | **[1.921, 12.700]** | **0.009** |
| **Model 2** | **7.328** | **[1.793, 12.864]** | **0.010** |
| **Model 3** | **6.351** | **[0.223, 12.480]** | **0.042** |
| *Hydroxycinnamic Acids* |  |  |  |
| Model 1 | -4.179 | [-8.475, 0.117] | 0.056 |
| Model 2 | -4.150 | [-8.572, 0.272] | 0.065 |
| Model 3 | -2.853 | [-7.580, 1.875] | 0.23 |
| *Verbal IQ* |  |  |  |
| *Flavones* |  |  |  |
| **Model 1** | **9.186** | **[3.570, 14.801]** | **0.002** |
| **Model 2** | **9.201** | **[3.448, 14.954]** | **0.002** |
| **Model 3** | **10.240** | **[3.870, 16.610]** | **0.002** |
| *Lignans* |  |  |  |
| **Model 1** | **6.433** | **[1.520, 11.347]** | **0.011** |
| **Model 2** | **6.366** | **[1.327, 11.405]** | **0.014** |
| **Model 3** | **6.769** | **[1.621, 11.917]** | **0.011** |

Estimates represent the association between prenatal polyphenol intake (averaged across pregnancy) and child cognitive outcomes (age 8 years) using linear regression models. Polyphenol intakes are energy-adjusted and log-transformed.
Model 1: Adjusted for maternal IQ and breastfeeding duration.
Model 2: Adjusted for the covariates in Model 2 plus maternal pre-pregnancy BMI, gestational age, and infant sex.
Model 3: Adjusted for the covariates in Model 2 plus child polyphenol intake using the same polyphenol measure as the prenatal polyphenol predictor.
Model 1 and 2, N = 73; Model 3, N = 71.

## Discussion

To our knowledge, this study is the first to demonstrate that prenatal intakes of specific (poly)phenol subclasses (flavones, lignans) are associated with increased language and IQ scores in school-age children. Specific (poly)phenol subclasses associating positively with child cognitive outcomes (flavones, lignans), while others associated negatively (flavonols, hydroxybenzoic acids), which may reflect contributions of (poly)phenol intakes that are specific to particular food sources co-consumed with other nutrients reflective of healthy (e.g., vitamin C, vitamin A, dietary fiber) or unhealthy plant-based foods (e.g., high sodium, added sugars, high saturated fat)^36, 51^. This study also characterized (poly)phenol intakes in pregnant women and their children in a longitudinal cohort to estimate total, class-level, and subclass-level (poly)phenol intakes. To our knowledge, this is the most comprehensive assessment of prenatal and child (poly)phenol intakes within a single cohort to date. There were 15 (poly)phenol subclasses consumed by mothers and their children, 9 of which had intakes greater than 10 mg/d during pregnancy and 5 mg/d in children. The major (poly)phenol in both prenatal and children’s diets was flavonoids, followed by phenolic acids, although children consumed more flavonoids and less phenolic acids. Major food sources of (poly)phenols consumed during pregnancy included coffee, blueberry, and okra, while major food sources for children’s intakes were okra, beans, plum, orange juice, and blueberry.

### (Poly)phenol intakes during pregnancy

Average (poly)phenol intakes were 716 ± 383 mg/d during pregnancy, which are within the ranges of median and mean intakes reported in international cohorts (320-3,367 mg/d)^24–27^, but are lower than population-level estimates in the U.S. (NHANES 2007-2016) for women (mean 1,545 mg/d) and adults age 20-39 years (mean 1,254 mg/d)^28^. Major (poly)phenol classes contributing to intake during pregnancy were phenolic acids and flavonoids, which is consistent with other pregnancy cohorts^25–27^ and population-level estimates in the U.S.^28^. However, the proportion of intake coming from phenolic acids (42%) and flavonoids (53%) differed. The contribution of phenolic acids to prenatal (poly)phenol intakes in our cohort was higher than other pregnancy cohorts in China (17% and 19%)^25, 26^ and Spain (22%)^27^ and lower compared to U.S. population-level estimates in adults (60%)^28^. The contribution of flavonoids to prenatal (poly)phenol intakes was lower compared to pregnant women in China (63% and 74%)^25, 26^, but higher than pregnant women in Spain (25-26%)^27^ and U.S. population-level estimates in adults (23%)^28^. Differences in intake may reflect heterogeneity in methodological approaches to collecting dietary intakes (e.g., 24-hour recall, food frequency questionnaire, 3-day food record)^24, 25, 27^, databases used for estimating (poly)phenol intakes (e.g., Phenol-Explorer with or without supplementation of content values from USDA flavonoid)^25–27^, or true differences in dietary intake as a result of sociodemographic factors (e.g., cultural or geographical dietary pattern differences) and life stage (e.g., pregnant women vs. non-pregnant adults).

Major food sources contributing to total (poly)phenol intakes during pregnancy were coffee, blueberry, and okra. Although prior studies have identified fruits, vegetables, and nuts as major food groups contributing to prenatal (poly)phenol intakes^25, 26^, only one study to date has characterized the major sources of prenatal (poly)phenol intakes at the level of individual food items^24^. Herbal and fruit teas and oranges were major contributors to total (poly)phenol intakes^24^. While these food items were not identified as major contributors to total prenatal (poly)phenol intakes in our cohort, we demonstrated that herbal teas are a major contributor to intakes of flavones, while oranges are a major source of flavanones. To our knowledge, our study is the first to characterize major food sources of prenatal (poly)phenol intakes at the class and subclass level for individual food items.

### (Poly)phenol intakes in school-age children

Mean total (poly)phenol intakes in children were similar at age 5 (395 ± 287 mg/d) and age 8 (411 ± 262 mg/d). Total (poly)phenol intakes for the children in our cohort were similar, but modestly higher, compared to children in Italy (8-10 years; mean 226 mg/d)^31^ and the UK (4-10 years; mean 389 mg/d)^33^, but lower compared to children in Iran (4-12 years; mean 1,641 mg/d)^30^. The high intakes reported for children in Iran is likely due to the methodology used (total (poly)phenol intake estimates based on Folin-Ciocalteu assay)^30^, whereas our estimates are based on the sum of individual (poly)phenol compounds measured by chromatographic methods.

It has been previously reported that estimating total (poly)phenol intakes using content values based on the Folin-Ciocalteu assay can overestimate intakes by 3-4 times^27, 31^. To our knowledge, the only subclass-level estimates of (poly)phenol intakes in children is for anthocyanin intakes derived from infant and toddler food intakes in Germany (3 months to 3 years; mean 0-4 mg/d with maximum ∼4 mg/d at 1.5-3 years of age)^34^. These intakes are lower compared to children in our cohort (20 mg/d at age 5; 22 mg/d at age 8), which likely reflects differences in dietary intake associated with child age. The major contributors to total (poly)phenol intakes in the children in our cohort were flavonoids (5 years: 66; 8 years: 69%) and phenolic acids (5 years: 29%; 8 years: 27%), which is similar to intakes in cohorts of healthy children in Italy (flavonoids: 76-80%; phenolic acids: 19-21% phenolic acids)^31^ and the UK (flavonoids: 80%; phenolic acids: 20%)^33^.

There are very few prior reports of major food sources contributing to (poly)phenol intakes in children^31, 33, 34^. Our study provides novel insights into (poly)phenol intakes in children in that we are, to our knowledge, the first to report major sources of (poly)phenol intakes at the level of individual food items at distinct ages (5 and 8 years) in school-age children in a U.S.-based cohort. Prior reports have focused on specific (poly)phenol subclasses (anthocyanins) in infants and toddlers^34^, broad age ranges (e.g., 4-10 years^33^, 8-10 years^31^), and, in some cases, lack food-level granularity^33^. Major sources of anthocyanins identified in children at age 3 years in Germany included strawberry and blueberry^33^, which we also reported as major contributors to anthocyanin intakes at age 5 and 8 in our cohort, in addition to other berries, including blackberry and sweet cherry. Compared to prior reports in school-age children from Italy^31^ and the UK, similarities in major food sources contributing to children’s (poly)phenol intakes include flavan-3-ols (apple^31^, tea^33^), flavanones (orange juice)^31^, flavones (orange juice, wheat)^31^, anthocyanins (strawberry)^31^, stilbenes (strawberry)^31^, and hydroxybenzoic acids (strawberry^31^, tea^31, 33^). In other cases, such as for total (poly)phenol intakes, flavonols, isoflavones, and hydroxycinnamic acids, differences in major food sources were observed between our cohort and prior reports^31, 33^. For example, we demonstrated that okra, tomato sauce, beans, and black tea were major sources of flavonol intakes for children in our cohort, while spinach was a major source of flavonols for children in Italy^31^. Similarly, several previously reported food sources of children’s hydroxycinnamic acids (apple, fruit juice, potatoes, tea)^31, 33^ were not reported as major food sources in our cohort. This may be due to the heterogeneity in (poly)phenol diversity for major food sources identified in prior studies and in our cohort^52^.

### Prenatal (poly)phenol intakes and offspring neurodevelopmental outcomes

Prior studies evaluating potential impacts of prenatal (poly)phenol intakes on offspring developmental outcomes are scarce, including two pilot RCTs^53, 54^ and one longitudinal birth cohort in Spain (GENEIDA project)^27^. The pilot RCTs tested the impact of a prenatal pomegranate juice intervention on neonatal brain structure in infants with intrauterine growth restriction, demonstrating lower risk of brain injury associated with increasing prenatal (poly)phenol intake derived from pomegranate juice^53, 54^. The cohort in Spain found associations between specific (poly)phenol subclasses (e.g., phenolic acids, stilbenes, anthocyanins) with birth characteristics including head circumference, height, and weight^27^. To our knowledge, our study is the first to demonstrate associations between prenatal (poly)phenol intakes and long-term neurodevelopmental outcomes. Similar to the cohort in Spain, we also found directionality of associations to be (poly)phenol-specific. Observations that specific (poly)phenols associate positively with child cognitive outcomes (flavones, lignans), while others associate negatively (flavonols, hydroxybenzoic acids) may reflect dietary sources and overall dietary patterns from which these (poly)phenols are derived. For example, in our cohort major food sources of flavones and lignans included orange juice (11% and 6% of flavone and lignan intakes, respectively), herbal tea (5% of flavone intakes), and kale (3% of lignan intakes), while hydroxybenzoic acids and flavonols were derived primarily from okra (45% of flavonol intakes), black tea (7% and 11% of hydroxybenzoic acid and flavonol intakes, respectively), tomato sauce (7% of flavonol intakes), and cranberry (9% of hydroxybenzoic acid intakes). Therefore, flavones and lignans may reflect higher intakes of foods and dishes that provide phytochemicals (including (poly)phenols), dietary fiber, and other micronutrients (e.g., vitamin C, vitamin A). Prenatal intakes of hydroxybenzoic acids and flavonols may reflect a less healthy profile of plant-based food intake. For example, okra was consumed cooked, cooked with breading, fried, or pickled. The three least healthy forms of okra intake (cooked with breading, fried, and pickled) represented 87% of okra-derived calorie intake. Nearly all caloric intake derived from black tea (97%) was consumed as sweetened black tea. Caloric intake derived from tomato sauce products included barbecue and steak sauces (34%), canned tomato products (e.g., tomato puree; 21%), pizza and marinara sauces (20%), spaghetti sauce (15%), and ketchup (11%), with 25% of intake derived from fast food meals. Intakes of okra and black tea, in particular, likely reflect dietary patterns typical of populations in the south and may not be reflective of intake patterns across the U.S. In addition, the modest effect sizes of the associations with language scores, a 7.5-point reduction in scores would require increasing hydroxybenzoic acid intake by ∼2,300 mg/d or flavonol intake by ∼6,700 mg/d. This corresponds to dietary intakes beyond what could feasibly be consumed through typical dietary intake (e.g., > 10 cups of black tea, > 2 cups of tomato sauce products, > 3 cups of dried cranberries, or > 10 cups of okra). Therefore, these foods are unlikely to have a clinically meaningful impact on child language development outcomes.

Our results are consistent with prior cohort studies in Canada (CHILD cohort)^35^, China^37^, the UK (ALSPAC cohort)^55^, and Japan (TMM BirThree Cohort Study)^36^ demonstrating that healthy (poly)phenol-rich foods and diets associate with improved cognitive outcomes in children. In the CHILD cohort in Canada, prenatal fruit intakes associated with higher cognitive scores at age 1 year^35^. In the cohort in China, increased Mediterannean diet adherence during pregnancy associated with reduced risk of communication development delay at age 1 year^37^. The ALSPAC cohort demonstrated that prenatal dietary patterns characterized by fruit and vegetable intake compared to meat and potatoes or white bread and coffee associated with higher child IQ scores at age 8 years^55^. The TMM BirThree Cohort Study found that increased prenatal fruit and vegetable intakes was associated with reduced risk of developmental delays at age 2 years^36^. Despite scarcity of studies adjusting for postnatal and child dietary intakes, these associations appear to be robust to adjustment for breastfeeding^36, 37^ and child dietary intake^36^. Our study expands on this by demonstrating that flavones and lignans provided by similar dietary profiles are associated with improvements in language skills and IQ scores in school-age children. These associations are also robust to adjustment for child diet covariates, including breastfeeding duration and child (poly)phenol intakes at age 5 and 8 years. An increase of 7.5 points (0.5 SD) is considered a modest, statistically significant effect size for cognitive outcomes^56^. In fully adjusted models, we demonstrated that this effect size would be achievable for improving child language skills and IQ scores by increasing prenatal flavone intakes by 458-552 mg/d (1/2 to 3/4 cup of coffee) and 52-62 mg/d (1/4 cup of coffee or 2-2.5 cups of herbal tea), respectively. This effect size is also achievable for improving verbal IQ scores by increasing prenatal lignan intakes by 255 mg/d (1/4 cup of coffee or 1.5 cups of kale). To date, only one study (TMM BirThree Cohort Study) has identified that associations between prenatal intakes of healthy plant-based foods (fruits and vegetables) and offspring cognitive outcomes are linked to intakes of specific dietary components (vitamin C, vitamin K, folate, β-carotene)^36^. Our study expands these dietary components to include specific subclasses of (poly)phenols (flavones, lignans). Therefore, future studies are warranted to delineate the contribution of (poly)phenols to these associations amongst other co-consumed nutrients such as vitamin K, vitamin C, vitamin A, carotenoids, folate, and dietary fiber.

Additionally, our results demonstrate domain-specific patterns in associations between prenatal (poly)phenol intakes and child cognitive outcomes, with prenatal flavone intakes predicting significant improvements in language skills and IQ scores and lignan intakes predicting higher IQ scores in school-age children, while no significant associations were observed for academic achievement scores. This is consistent with most prior studies that have linked prenatal intakes of (poly)phenol-rich foods and dietary patterns with improvements in specific neurodevelopmental domains in children, including cognition^35^, communication^37^, and IQ scores^55^. Only the TMM BirThree Cohort Study reported broader cross-domain associations between prenatal fruit and vegetable intakes and reducing risk of developmental delays in communication, motor skills, problem solving, and personal-social skills at age 2 years^36^. Differences across studies may be due to differences in dietary assessment methodologies and time points, child age, cognitive assessment instruments used, population characteristics, and adjustment for child diet covariates.

### Potential mechanistic pathways linking prenatal (poly)phenol intakes to child cognitive health

Compared to studies linking (poly)phenol intake to cognitive health outcomes in adults, identification of potential roles for (poly)phenol intakes to improve cognitive health outcomes in children is relatively recent. For example, Ross et al.^53^ and Matthews et al.^54^ demonstrated potential neuroprotective effects for infants whose mothers increased their (poly)phenol intakes during pregnancy through a pomegranate juice intervention. Consistent with this paucity of evidence linking early-life (poly)phenol exposures to offspring neurodevelopmental and cognitive function outcomes, the mechanism of action for potential neurodevelopmental impacts of early-life (poly)phenol intakes are poorly understood. In the context of prenatal exposures, possible mechanisms may involve the direct transfer of (poly)phenol metabolites to the developing fetal brain. Although transfer of (poly)phenol metabolites from maternal circulation to the developing fetus have yet to be evaluated in humans, rodent models demonstrate that (poly)phenol transport across the placenta is low but does indeed occur^57, 58^. A framework for such clinical studies to be conducted in the future could be drawn from the recent Lutein and Zeaxanthin in Pregnancy (L-ZIP) study, which evaluated efficacy of prenatal carotenoid supplementation on carotenoid status of mothers and their infants and explored changes in ocular development^59^. While (poly)phenol transfer has yet to be evaluated during pregnancy, transfer of flavan-3-ol metabolites to plasma and brain have been demonstrated in a preclinical piglet model reflective of intakes in children at 1.5-3 years of age^60^. However, inconsistencies in the dose-dependency of this transfer have been observed^60, 61^, with preferential transfer of specific metabolites derived from gut microbial metabolism of dietary (poly)phenols^62, 63^. Mechanisms that have been proposed for neuroprotection by (poly)phenols include indirect effects related to reductions in nitric oxide and inflammation^60^, which may occur locally in the developing brain as demonstrated through preclinical cell culture models^64^. Consistent with this postulated mechanism, *in vitro* studies have demonstrated that phenolic sulfates derived from microbial metabolism of (poly)phenols in berries cross the blood brain barrier endothelial cells and modulate microglia-mediated neuroinflammation^64^. Effects of prenatal (poly)phenol intakes on development of fetal brain structure, connectivity, and function have also been suggested^53, 54^.

It is also possible that potential improvements in child neurodevelopmental outcomes associated with prenatal (poly)phenol exposures may be mediated by improvements in maternal cardiometabolic health status throughout pregnancy. It is well-established that maternal cardiometabolic health has deleterious programming impacts on later childhood cognitive health outcomes, including IQ scores^65^, academic skills^66^, executive function^67^, mental and psychomotor development^68^, neurodevelopmental disorders^69, 70^, developmental delays^70, 71^, and emotional and behavioral challenges^70^. It has been demonstrated that increasing (poly)phenol intakes in adults leads to improvements in cardiometabolic health outcomes^72–77^. A recent cohort study in China (Tongjui Birth Cohort) has extended these observations to cardiometabolic outcomes during pregnancy outcomes, demonstrating that prenatal (poly)phenol intakes are associated with reduced risks for gestational diabetes^25^ and hypertension^26^. These results suggest a plausible indirect mechanistic pathway for prenatal (poly)phenol exposures involving mediation by maternal cardiometabolic health status during pregnancy. Future studies are needed to evaluate transfer and dose-dependency of (poly)phenol metabolites in maternal circulation, availability to the fetus, and neonatal circulation. In addition, further work is needed to delineate specific neurodevelopmental pathways, exposure timing, dose, and contribution of sustained or accumulated exposures across pregnancy, breastfeeding, and early child feeding.

### Strengths and Limitations

Strengths of our study include the estimation of total (poly)phenol intakes, and class- and subclass-level intakes across pregnancy and in school-age children. This study provides an in-depth methodological workflow for extracting these intakes from Phenol-Explorer, facilitating its application for future analyses. Our study based (poly)phenol intake estimates on the chromatographic data in Phenol-Explorer, which allows quantitation of various (poly)phenol classes and subclasses, unlike the Folin-Ciocalteu assay that provides data only for total (poly)phenols^78^ or other databases (e.g., USDA Flavonoid) that provide data on a specific (poly)phenol class^79^. Other strengths include longitudinal dietary assessments across multiple time points throughout pregnancy and childhood, allowing for more robust estimates of (poly)phenol intakes during pregnancy and childhood. In comparison, several prior studies have relied on only one or two time points during pregnancy^25–27^, limiting their ability to account for changes in dietary intakes across pregnancy. In addition, child cognitive assessments were evaluated at multiple time points across multiple domains enabling evaluation of longitudinal trajectories of neuropsychological function. Finally, our cohort includes data collection across pregnancy, infancy, and childhood, which allows adjustment for critical child diet covariates, including breastfeeding duration and child (poly)phenol intakes.

Limitations of our study include the challenge of comparing results with previous studies due to differences in methodologies of assessing dietary intake and workflows for extracting (poly)phenol intake estimates. In prior studies that estimated prenatal (poly)phenol intakes, food frequency questionnaires were the predominant methodology^24–27^, with a few studies utilizing 24-hour recalls^24, 27^ and 3-day food records^24^. While systematic misreporting of dietary intakes occurs across all forms of self-reported dietary intake^80^, food records are considered to provide more accurate estimations because they are prospective and do not rely on the participant’s memory^81^. In addition, there is considerable heterogeneity in approaches utilized to estimate (poly)phenol intakes during pregnancy including USDA Flavonoid and Phenol-Explorer databases with and without supplementation using external data sources^24–27^. Our detailed workflow enables reproducibility in future studies, even though our methodology differs from previous work. In addition, this study could facilitate future exploration of associations between specific types of (poly)phenols (at the individual compound level) and maternal-child health outcomes. Another limitation of our study is that there may be inaccuracies in quantitative (poly)phenol estimates, which is an inherent limitation of methodology that relies on databases with quantitative estimates of food (poly)phenol content. All such databases (e.g., Phenol-Explorer, USDA Flavonoid) rely on literature reports of quantitative values in foods and therefore reflect content of that particular food sample. However, agronomic factors (e.g., season^82^, growing conditions^83, 84^, cultivar/accession^82, 85^, stage of ripening^86^), storage conditions^87^, and food formulation (e.g., vitamin-enhanced water or carbonated beverages may be formulated with (poly)phenol-rich concentrates or extracts)^88^ and preparation/processing^89^ will introduce inaccuracies in actual (poly)phenol intakes. Ideally, (poly)phenol concentrations would be measured directly in foods consumed by participants. However, in most longitudinal clinical studies this is not feasible due, in part, to analytical and sample collection challenges, including both financial cost and participant burden, associated with accurately quantifying individual (poly)phenols in every food item consumed (e.g., 2,229 unique (poly)phenol-containing food items in our cohort). Therefore, at present, inferring (poly)phenol intakes using databases such as Phenol-Explorer and USDA Flavonoid is considered to provide the most complete description of (poly)phenol intakes^43^ and has demonstrated utility for exploring potential impacts on maternal and child health outcomes^25–27^. Another limitation of our study is the lack of data for both mothers and their children on (poly)phenol metabolite concentrations in plasma or biospecimens collected at birth (e.g., cord blood, placenta). Future studies are needed to explore these metabolites in relation to neurodevelopmental outcomes since these metabolites have been associated with measures of cognitive function in adult populations^90^. In addition, child cognitive development was not the primary outcome of the parent cohort or follow-up study and, therefore, the analysis may be underpowered to detect modest associations. Findings of our study also need to be replicated in larger, more diverse cohorts. Finally, due to the observational design of our study, causality cannot be established between prenatal (poly)phenol intakes and child cognitive outcomes. However, this study provides a basis for future efforts to evaluate efficacy of (poly)phenol-rich interventions during pregnancy for improving offspring neurodevelopmental outcomes.

## Conclusions

Our study provides a valuable resource for future research due to the depth of (poly)phenol characterization in both mothers during pregnancy and their children, which is unique in the field of (poly)phenol research and provides a framework for estimating (poly)phenol intakes in other studies. This study demonstrated associations between prenatal intakes of specific (poly)phenols and long-term cognitive health outcomes during childhood for the first time, providing a basis for development of more targeted nutrition interventions and future studies to establish bioavailability, pharmacokinetics, and transfer of (poly)phenol metabolites throughout pregnancy. We also showed that the directionality of associations is subclass-specific and may reflect intakes of particular plant-based foods or co-consumption of specific nutrients or dietary components. These findings will inform future studies exploring the contribution of (poly)phenols within the context of broader diet and food composition. Overall, the results of this study strengthen existing evidence suggesting that early-life (poly)phenol exposures impact offspring neurodevelopmental outcomes and point to the need for delineating potential mechanisms responsible for these associations.

## Abbreviations

BMI: Body mass index
CELF-5: Clinical Evaluation of Language Fundamentals, 5^th^ edition
DHA: Docosahexaenoic acid
GWG: Gestational weight gain
IQ: Intelligence quotient
L-ZIP: Lutein and Zeaxanthin in Pregnancy
NASEM: National Academy of Sciences, Engineering, and Medicine
NDSR: Nutrition Data System for Research
WASI-II: Wechsler Abbreviated Scale of Intelligence, 2^nd^ edition
WRAT4: Wide Range Achievement Test, 4^th^ edition

## Acknowledgments

The authors are grateful to the children and families who participated in this study and to the clinical core team for their assistance in data collection. The authors acknowledge the Maternal and Early Nutrition and Development (MEND) core team members of the Arkansas Children’s Nutrition Center for their support in conducting the parent study.

## Author contributions

**C Fiecke:** Conceptualization, Methodology, Formal analysis, Writing - Original Draft, Visualization, Writing - Review & Editing. **J Bellando:** Data curation, Investigation, Methodology, Writing - Review & Editing. **M Whiteside:** Data curation, Investigation, Methodology, Writing - Review & Editing. **C Slaton:** Data curation, Investigation, Methodology, Writing - Review & Editing. **M Bimali:** Formal analysis, Writing - Review & Editing. **M Bao:** Formal analysis, Writing - Review & Editing. **DK Williams:** Formal analysis, Writing - Review & Editing. **E van der Plas:** Writing - Review & Editing. **CD Kay:** Writing - Review & Editing. **A Andres:** Conceptualization, Formal analysis, Methodology, Funding acquisition, Writing - Review & Editing. All authors read and approved the final manuscript.

## Data Availability

Deidentified individual participant data, code book, and analytic code will be made available upon reasonable request.

## Declaration of Generative AI and AI-assisted technologies in the writing process

The author(s) declare that no generative AI or AI-assisted technologies were used in the writing of this manuscript.

